# Smoking and the risk of COVID-19 infection in the UK Biobank Prospective Study

**DOI:** 10.1101/2020.05.05.20092445

**Authors:** Eo Rin Cho, Prabhat Jha, Arthur S. Slutsky

## Abstract

Several studies suggest a lower prevalence of smoking than expected among adults with coronavirus disease (COVID-19). We conducted logistic regression analyses of the UK Biobank prospective study of 0.5 million adults followed for an average of 11 years. Compared to women, men were more likely to be tested and to test positive. In sex-stratified analyses, current smokers had higher adjusted Odds Ratios (OR) for being tested (male OR 1.60, 95%CI 1.32-1.95 and female OR 1.50,1.21-.1.86). Current smokers were more slightly more likely than never smokers to test positive for COVID-19. Further examination of smoking as a risk factor for COVID-19 is required. These must take into account reverse causality, where smokers quit to avoid disease as well as prior diseases.

## Introduction

Several studies suggest a lower prevalence of smoking than expected among adults with coronavirus disease (COVID-19).^1^

Proper assessment of the risks of smoking and COVID-19 is best done in prospective studies. We analyzed smoking, obesity and other risk factors in the UK Biobank, a prospective study of 502,506 adults age 40-86 years followed for an average of 11.3 years.^2^ Between March 16-April 26, 2,237 participants were tested for COVID-19 infection. 908 (40.6%) tested positive, suggesting that testing was of patients at high-risk of COVID-19. However, the smoking prevalence at enrollment was slightly lower in those testing positive (151/908; 16.6%) than testing negative (232/1329; 17.5%). But this was mostly a spurious result.

We conducted logistic regression analyses controlling for age (at time of COVID-19 testing), and the following variables at enrollment: smoking (current or quit < 5 years earlier; former, meaning quit >5 years earlier; never); body mass index (BMI; weight/height^2^ of <27, 27-<30, ≥30); and self-reported diabetes or hypertension. We excluded those who had developed vascular, respiratory or neoplastic diseases by March 2020. Compared to women, men were more likely to be tested (adjusted Odds ratio [OR]=1.13; 95% CI 1.01-1.26) and to test positive (OR=1.35;1.07-1.69). In sex-stratified analyses, current smokers had higher ORs for being tested (men's OR=1.60; 1.32-1.95; women's OR=1.50; 1.21-1.86). Former smokers, those with BMI >27, a history of hypertension or, among men, a history of diabetes were also more likely to be tested (Table).

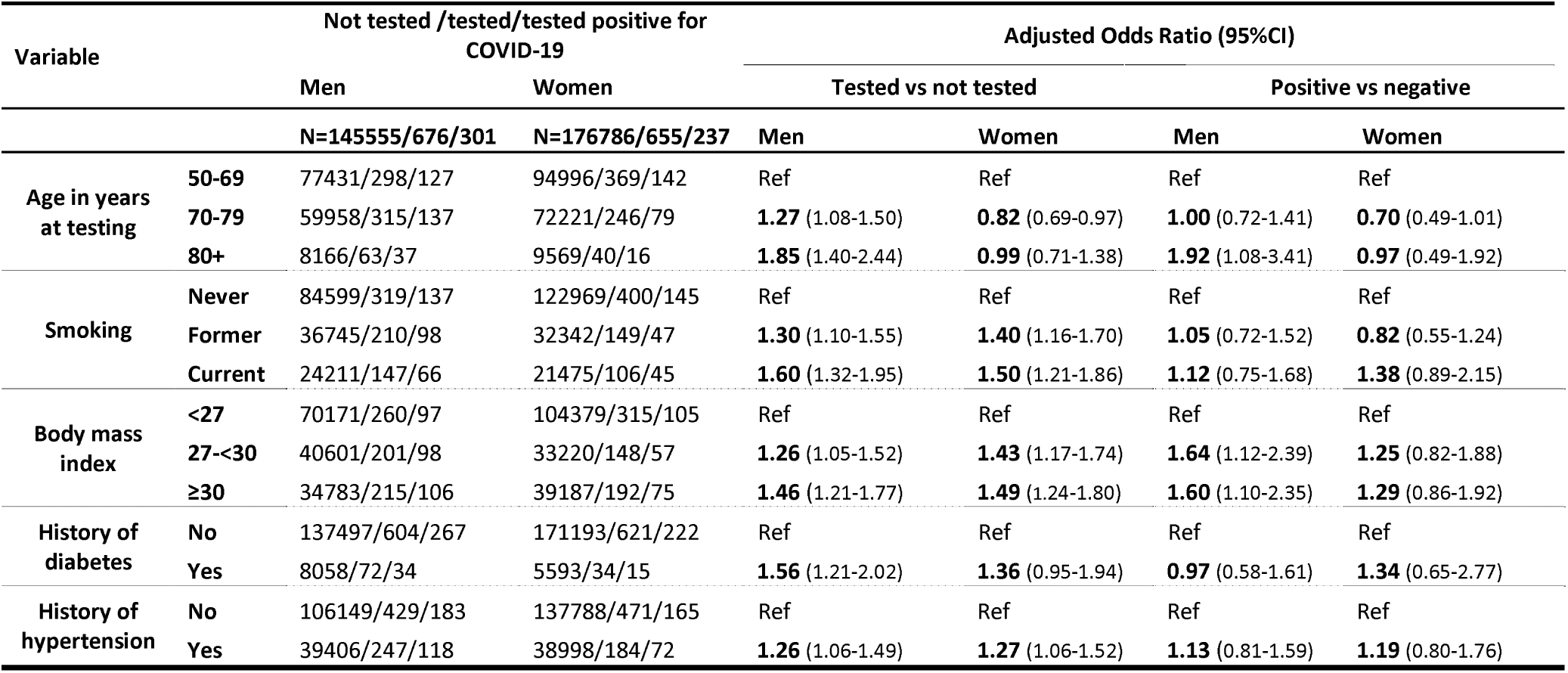
Table Determinants of being tested for COVID-19 and of testing positive among UK Biobank participants followed prospectively for 11.3 years. Notes: We used age at the time of COVID-19 testing (Mar 16-Apr 26, 2020) and data at enrollment on smoking status, BMI, diabetes and hypertension. We excluded those who by the time of follow-up had died (n=20,283) or had developed cancer (34,418), vascular (23,348) or respiratory (50,053) disease, or were missing the following variables: BMI (2,053), smoking status (1,681), age quit smoking (42,638) or self-reported history of disease (4,360). Exclusions among 2,237 tested were as follows: developed cancer (201), vascular (197) or respiratory (259) diseases; missing BMI (22), smoking status (7), age quit smoking (168) or self-reported history of disease (52). Odds ratios adjusted for age, smoking status, BMI, and medical history of diabetes/hypertension. Results not excluding those whose age at quitting was unknown were similar, as was the comparison of ever smokers (i.e. current and former) versus ne/er smokers.

After adjusting for these variables, current smokers were slightly more likely than never smokers to test positive COVID-19 tests, but this was not statistically significant: men's OR=1.12; 0.75-1.68; women's OR=1.38; 0.89-2.15. Former smoking was similarly unassociated with a positive test. Only being >80 years of age or having a BMI >27 among men were significantly associated with a positive test. No variables were significantly associated with a positive test in women. Results were similar comparing ever versus never smokers.

Further examination of smoking as a risk factor for COVID-19 in other prospective studies is required. This must take into account not only reverse causality, where smokers quit to avoid disease^3^, but also prior diseases and co-morbidities including obesity, diabetes and hypertension, as each is associated with COVID-19 hospitalization or mortality.^4^ COVID-19 has thus far killed over 240,000 people mostly in high-income countries.^5^ That total reflects about 20 days of annual global tobacco deaths.^3^ Both epidemics should be considered seriously and measured appropriately.

COI Statement: AS is a co-founder of a company that has developed a nicotine replacement product; this company is working with Phillip Morris International. ERC and PJ report no potential conflict of interest relevant to this letter.

## Data Availability

Data from the UK Biobank are openly available.

